# Upper limb functional recovery in chronic stroke patients after COVID-19-interrupted rehabilitation: An observational study

**DOI:** 10.1101/2023.06.05.23290309

**Authors:** Daigo Sakamoto, Toyohiro Hamaguchi, Yasuhide Nakayama, Takuya Hada, Masahiro Abo

## Abstract

**Background:** Upper limb function of chronic stroke patients declined when outpatient rehabilitation was interrupted, and outings restricted, due to the severe acute respiratory syndrome coronavirus 2 (SARS-CoV-2) pandemic. In this study, we investigated whether these patients recovered upper limb function after resumption of outpatient rehabilitation.

**Methods:** In this observational study, 43 chronic stroke hemiplegic patients with impaired upper extremity function were scored for limb function via Fugl-Meyer Assessment of the Upper Extremity (FMA-UE), Action Research Arm Test (ARAT) after a structured interview, evaluation, and intervention. Scores at 6 months and 3 months before and 3 months after rehabilitation interruption were examined retrospectively, and scores immediately after resumption of care and at 3 and 6 months after resumption of care were examined prospectively. The amount of change for each time period and an analysis of covariance was performed with time as a factor and the change in FMA-UE and ARAT scores as dependent variables and by setting statistical significance at 5%.

**Results:** Time of evaluation significantly impacted total, part C, and part D of FMA-UE as well as total, pinch, and gross movement of ARAT. Post-hoc tests showed that the magnitude of change in limb function scores from immediately after resumption of rehabilitation to 3 months after resumption was significantly higher than the change from 3 months before to immediately after interruption for total, and part D of FMA-UE, and grip, and gross movement of ARAT (*p*<0.05).

**Conclusions:** The results suggest that upper limb functional decline in chronic stroke patients, caused by the SARS-CoV-2 pandemic-related therapy interruption and outing restrictions, was resolved after approximately 3 months of resumption of rehabilitation therapy. Our data can serve as reference standards for planning and evaluating treatment for chronic stroke patients with impaired upper limb function due to inactivity.

## Introduction

Measures implemented to control a pandemic of novel coronavirus infection (COVID-19) caused by severe acute respiratory syndrome coronavirus 2 (SARS-CoV-2) appear to have had a negative impact on the health of some people. COVID-19 spread rapidly after its outbreak in December 2019, and the World Health Organization declared it a pandemic [1]. Lockdowns were implemented around the world to prevent the spread of SARS-CoV-2, and a state of emergency was declared in Japan [2–4]. People were forced to change their lifestyles, as non-essential outings were restricted. People spent more time at home, and this led to longer sedentary hours and less physical activity during the day [5]. The decrease in physical activity in the elderly and patients receiving physiotherapy caused physical and mental harm [6, 7].

COVID-19 related interruption in care for patients with hemiplegia after chronic stroke it has been reported to result in worsening of upper limb motor function and subjective physical symptoms [8]. Post-stroke motor paralysis limits patient activities of daily living (ADL) and reduces their quality of life [9, 10]. Continuous rehabilitation is important to maintain and improve ADL and quality of life of stroke patients [11]. However, the SARS-CoV-2 pandemic interrupted care for chronic outpatients, and patients temporarily lost access to treatment and guidance from therapists [12]. This interruption showed patients and health care providers that patients risk losing physical functionality when rehabilitation is interrupted for a period of time [8]. If this temporary decline in physical functionality can be reversed with subsequent support, the importance of continued rehabilitation will be recognized.

The purpose of this study was to estimate the amount of recovery of upper limb function in chronic stroke patients after COVID-19 related interruption of outpatient care. In this study, chronic stroke patients we had previously studied [8] were re-investigated after a practice plan was developed and rehabilitation was resumed within the scope of usual practice. The upper extremity function of patients was restored within approximately 3 months after resumption of rehabilitation. The results of this study can be used as a reference for future planning of exercises as well as for devising ways to prevent the decline in upper limb function for chronic stroke patients.

## Materials and Methods

### Study design

This was an observational study on pre- and post-survey data without a control group.

### Participants

Our study included patients who had received outpatient occupational therapy for at least 6 months at the Department of Rehabilitation Medicine of the Jikei University Hospital between June 1, 2019, and May 31, 2020, were post-stroke hemiplegic patients for whom at least 6 months had passed since onset, were at least 20 years of age, and were patients whose outpatient rehabilitation was temporarily interrupted due to the spread of SARS-CoV-2 infection. Patients were excluded if they had a diagnosis of higher brain dysfunction, cognitive impairment, or a psychiatric disorder that would affect functional assessment measures and understanding of instructions during rehabilitation. After confirming that the patients met the eligibility criteria, we provided written and oral explanations of the study and requested their participation. Patients were recruited starting June 1, 2020. Those who agreed to participate were considered eligible for the study.

The minimum sample size was calculated to be 40 cases in total by using G*Power 3.1 software, with a goodness of fit test (F-test) and analysis of variance (for fixed effects, omnibus, one-way ANOVA), effect size f = 0.73, α = 0.05, power = 0.95, number of groups = 4, numerator df = 3, and partial η^2^ = 0.35.

### Survey periods and instruments

This study was conducted approximately 6 months before outpatient rehabilitation was interrupted (6 m before), approximately 3 months before outpatient rehabilitation was interrupted (3 m before), after interruption period (After IP), approximately 3 months after outpatient rehabilitation was resumed (3 m after), and approximately 6 months after outpatient rehabilitation was resumed (6 m after). The date on which outpatient occupational therapy was suspended due to the SARS-CoV-2 infection outbreak was April 1, 2020. At 6 m before and 3 m before, Fugl-Meyer assessment of the upper extremity (FMA-UE) and Action Research Arm Test (ARAT) scores were examined retrospectively from the medical records. The Medical records from 1 June 2019 to 31 May 2020 were collected. At After IP, 3 m after, and 6 m after, FMA-UE and ARAT scores were prospectively assessed (Fig 1).

**Fig 1.**
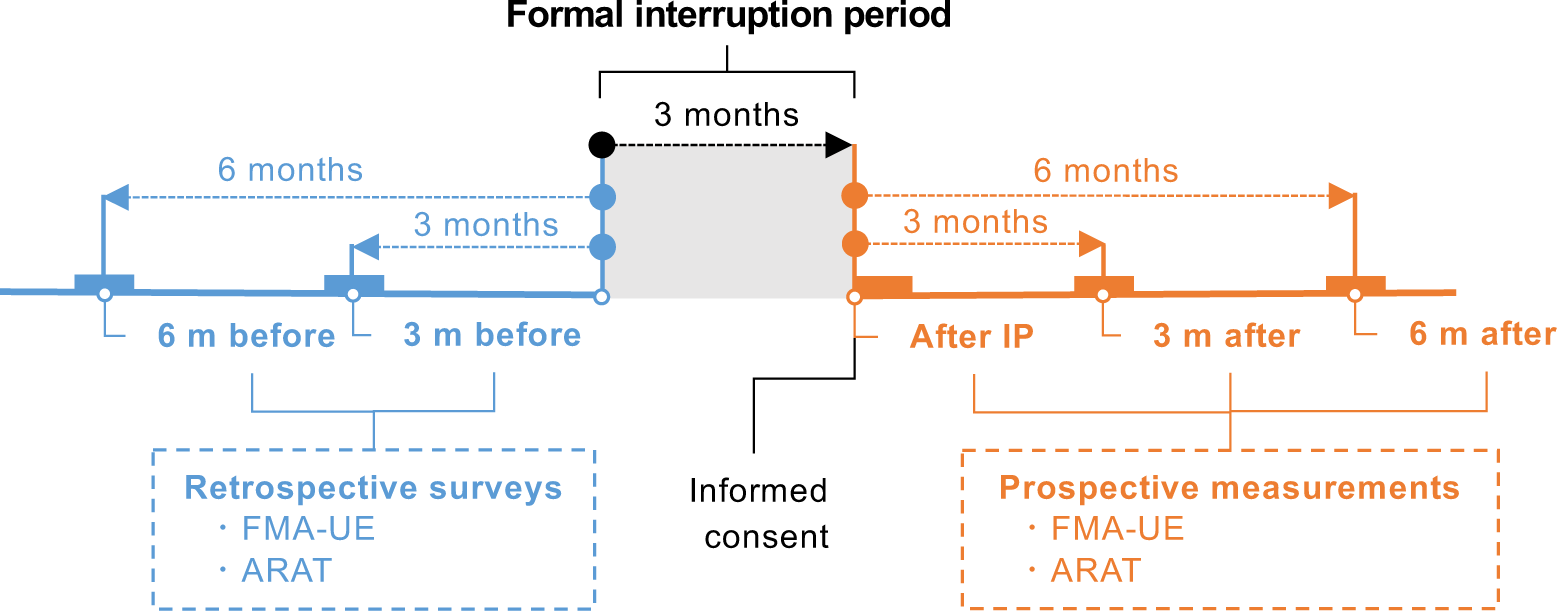
Schematic showing timing of limb function surveys in relation to the therapy interruption period.

6 m before, approximately 6 months before outpatient rehabilitation was interrupted; 3 m before, approximately 3 months before outpatient rehabilitation was interrupted; After IP, after interruption period; 3 m after, approximately 3 months after outpatient rehabilitation was resumed; 6 m after, approximately 6 months after outpatient rehabilitation was resumed; FMA-UE, Fugl-Meyer assessment of the upper extremity; ARAT, Action Research Arm Test.

### Occupational Therapy for Outpatients

Occupational therapists conducted structured interviews, assessments, and interventions within the usual scope of practice with patients who had resumed outpatient rehabilitation (Table First, occupational therapists interviewed patients about subjective symptoms that occurred during the 3-month period when outpatient rehabilitation was suspended and patients were not allowed to leave the house, using an originally designed questionnaire. The questionnaire was developed based on the International Classification of Functioning, Disability and Health (ICF) category [13] and consisted of questions regarding function, activity and participation (Appendix). The occupational therapist set clear treatment goals in consultation with the patient, considering the subjective symptoms that occurred in the patient. Next, the occupational therapist assessed the motor paralysis, joint range of motion, and muscle tone in the upper extremities and fingers of the patient. These assessments were performed during a traditional occupational therapy session. The FMA and ARAT scores of the patient that assess motor paralysis were compared to previous assessments to determine any changes in scores. The occupational therapist also interviewed the patient about the quality and frequency of use of ADL in the paralyzed upper extremity, and about the self-training of the patient at home.

The occupational therapist shared a practice plan with the patient that was developed in consideration of subjective symptoms that occurred in the patient and provided practice and instruction. The occupational therapists identified items with decreased ratings in the upper extremity functionality of the patient that were a high priority for treatment, described exercises to improve limb function, and guided the patient in self-training. The main kinds of exercises included joint range of motion exercises to improve joint contractures, stretching exercises for spastic muscles to decrease muscle tension, and upper limb function exercises to improve motor paralysis. Pamphlets containing the instructions were distributed to the patients. The instructions were modified or added to as the subjective symptoms and upper extremity functionality of the patients changed. In the second and subsequent sessions, the occupational therapist provided positive feedback to the patient on the performance of self-training and changes in limb function and movement to maintain and improve patient motivation.

### Main outcome

The change in FMA-UE scores was used to assess the primary outcome. The FMA is a comprehensive assessment battery that examines motor function, balance function, sensory function, joint range of motion, and joint pain in post-stroke hemiplegic patients [14, 15]. The items relating to motor function of the upper limbs can be used in excerpts. The FMA-UE has four sections: part A assesses shoulder, elbow, and forearm motion; part B assesses wrist motion; part C assesses finger motion; and part D assesses coordinated upper extremity motion. The upper extremity motor function items are scored on a 3-point ordinal scale with a maximum score of 66 points. The FMA-UE scores were classified according to the report of Woodbury et al. to determine the severity of motor paralysis [16].

### Secondary outcome

The change in ARAT scores was used to assess the secondary outcome. The ARAT is an upper extremity functional assessment developed based on the Upper Extremity Function Test [17]. The ARAT consists of grasp, grip, pinch, and gross movement items, and includes an object manipulation task. While the FMA-UE is an assessment method that captures patient function, the ARAT is reported to reflect patient activity [18]. The test is scored on a 4-point ordinal scale with a maximum score of 57.

### Participants characteristics

The characteristics of the subjects were investigated in terms of age, gender, height, weight, Body Mass Index (BMI), dominant hand, and Barthel Index (BI). The BI is a test to evaluate patient independence in ADL [19]. The BI consists of 10 items and is scored out of 100 points on a 4-point ordinal scale for the degree of assistance. If the patient is independent in all items, the score is 100 points, and if the patient requires full assistance in all items, the score is 0 points. As part of patient information, the type of stroke, the side of paralysis, the duration since the onset of stroke, whether or not botulinum treatment was given, the duration of discontinuation of outpatient rehabilitation, the duration at each evaluation point, the number of occupational therapy sessions per month, and the duration per session were investigated.

### Investigators

Investigations of upper limb motor function, ADL, and treatment of patients were conducted by seven occupational therapists working in the Jikei University Hospital and engaged in rehabilitation in the area of cerebrovascular disorders for more than 5 years.

### Statistical analysis

The total scores of FMA-UE and ARAT and the scores of the sub-items were calculated as the amount of change (delta) using the following equation to estimate their recovery.

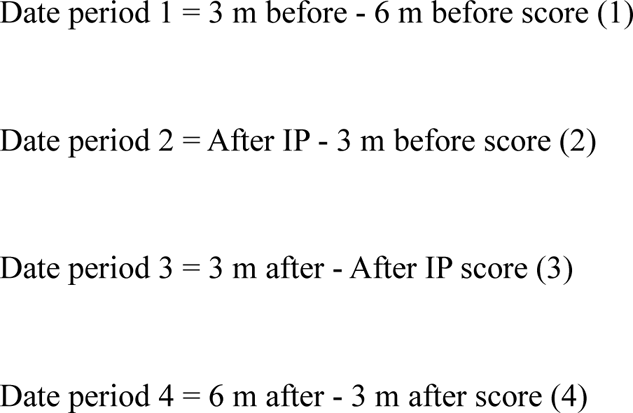

To test the hypothesis that resumption of outpatient rehabilitation improves upper limb motor function in patients with hemiplegia due to chronic stroke whose upper limb motor function had declined as a result of interrupted outpatient rehabilitation and refraining from going outside, we used time as a factor and change in FMA-UE and ARAT as dependent variables. An analysis of variance was conducted. The covariates were age, gender, BMI, time since onset, and the 6 m before score [20, 21]. JASP 0.16 software (University of Amsterdam Department of Psychology & Psychological Methods Unit, Amsterdam, the Netherlands) was used for the statistical analysis. A *p*-value of <0.05 was considered statistically significant.

### Ethical considerations

All patients who participated in the study gave written consent to participate in the study. The study was approved by the Ethics Committee of the Jikei University School of Medicine (Approval Number 24-295-7061). Patients were examined by a rehabilitation physician prior to evaluation and treatment by an occupational therapist.

Patients with a temperature of 37°C or higher, upper respiratory symptoms, loss of taste and smell symptoms, and other common cold symptoms at the time of examination were withdrawn from rehabilitation and laboratory measurements. To prevent transmission of infection to occupational therapists and patients, all patients washed their hands and disinfected with alcohol before entering the rehabilitation room. Patients wore masks, and occupational therapists wore masks, gowns, and rubber gloves.

## Results

Between June 1, 2019, and May 31, 2020, there were 81 patients with chronic stroke hemiplegia enrolled in our study who underwent at least 3 months of outpatient rehabilitation. Of the 81 patients, 4 patients younger than 20 years and 4 patients whose occupational therapy intervention was completed before the interruption of outpatient occupational therapy due to the spread of novel coronavirus infection were excluded. Of the 73 patients who met the study eligibility criteria, 21 patients who did not wish to resume outpatient occupational therapy and 3 patients who did not give consent to participate in the study were excluded, resulting in 49 patients who participated in the study. It has been reported by us previously that the upper limb functions of the patients deteriorated, and subjective physical symptoms occurred after approximately 3 months of interruption of outpatient occupational therapy [8]. In the present study, we followed up on these 49 patients. Two patients whose occupational therapy was terminated during the follow-up period and four patients who requested discontinuation of outpatient occupational therapy due to the re-emergence of novel coronavirus infection were excluded from the analysis. The total number of patients included in the final analysis of the present study was 43 (Fig 2). The characteristics of the patients included in the present analysis are shown in Table 2. The FMA-UE and ARAT scores during the study period are shown in Table 3.

**Fig 2.**
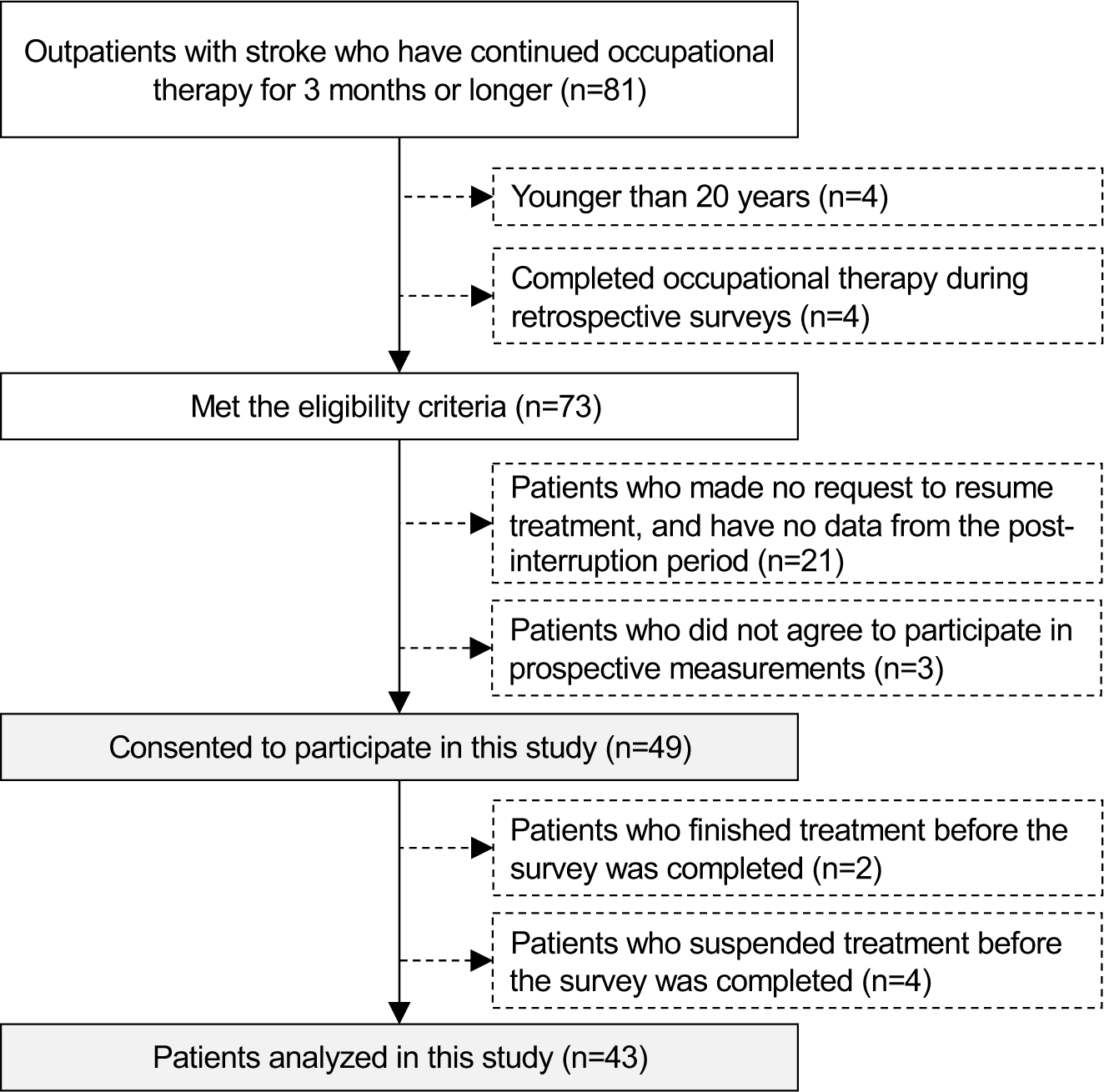
Flow chart showing patient inclusion criteria.

**Table 1.**
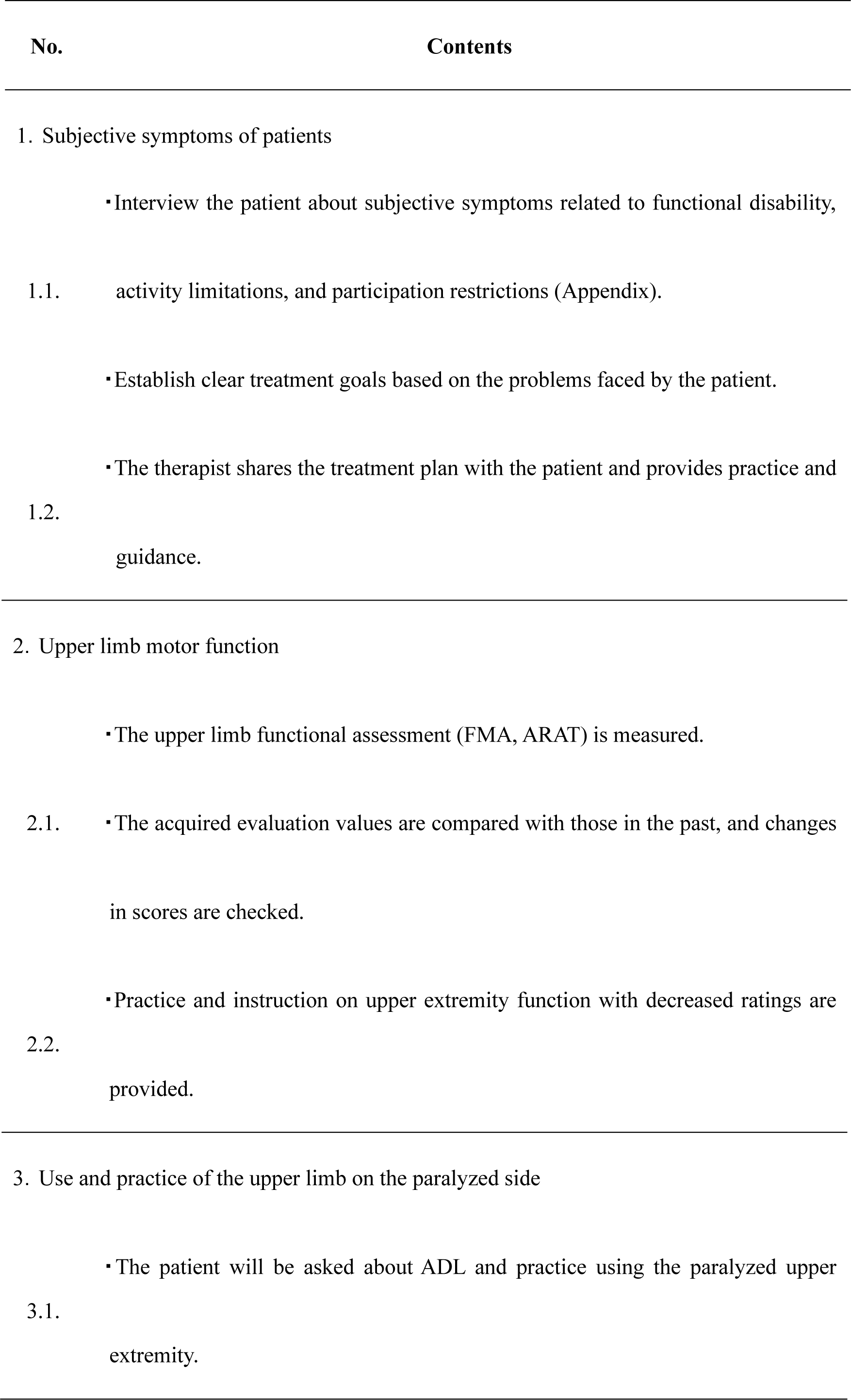
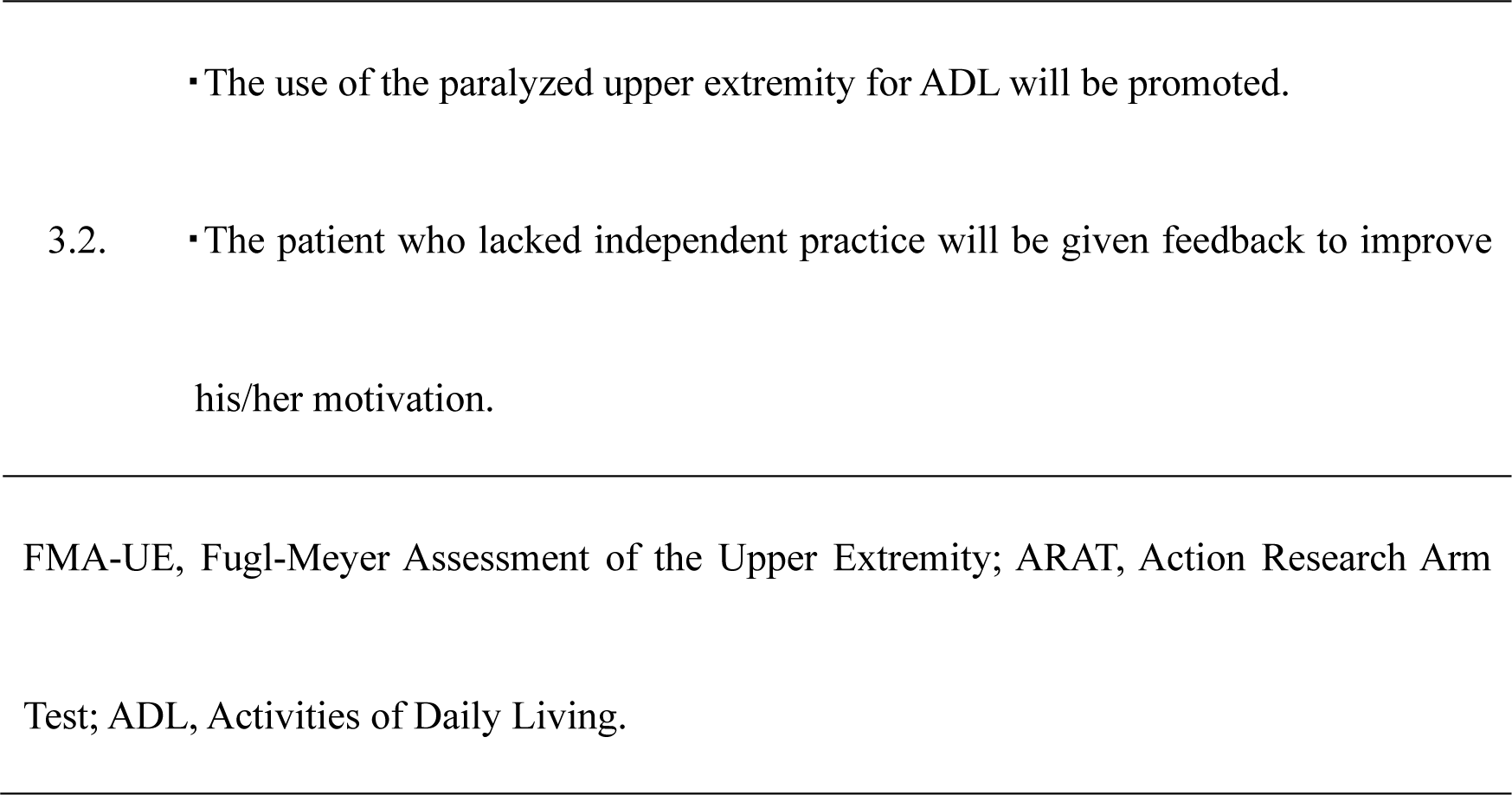
Structured interviews, assessments and interventions performed within the usual scope of practice for patients who resumed outpatient rehabilitation.

**Table 2.**
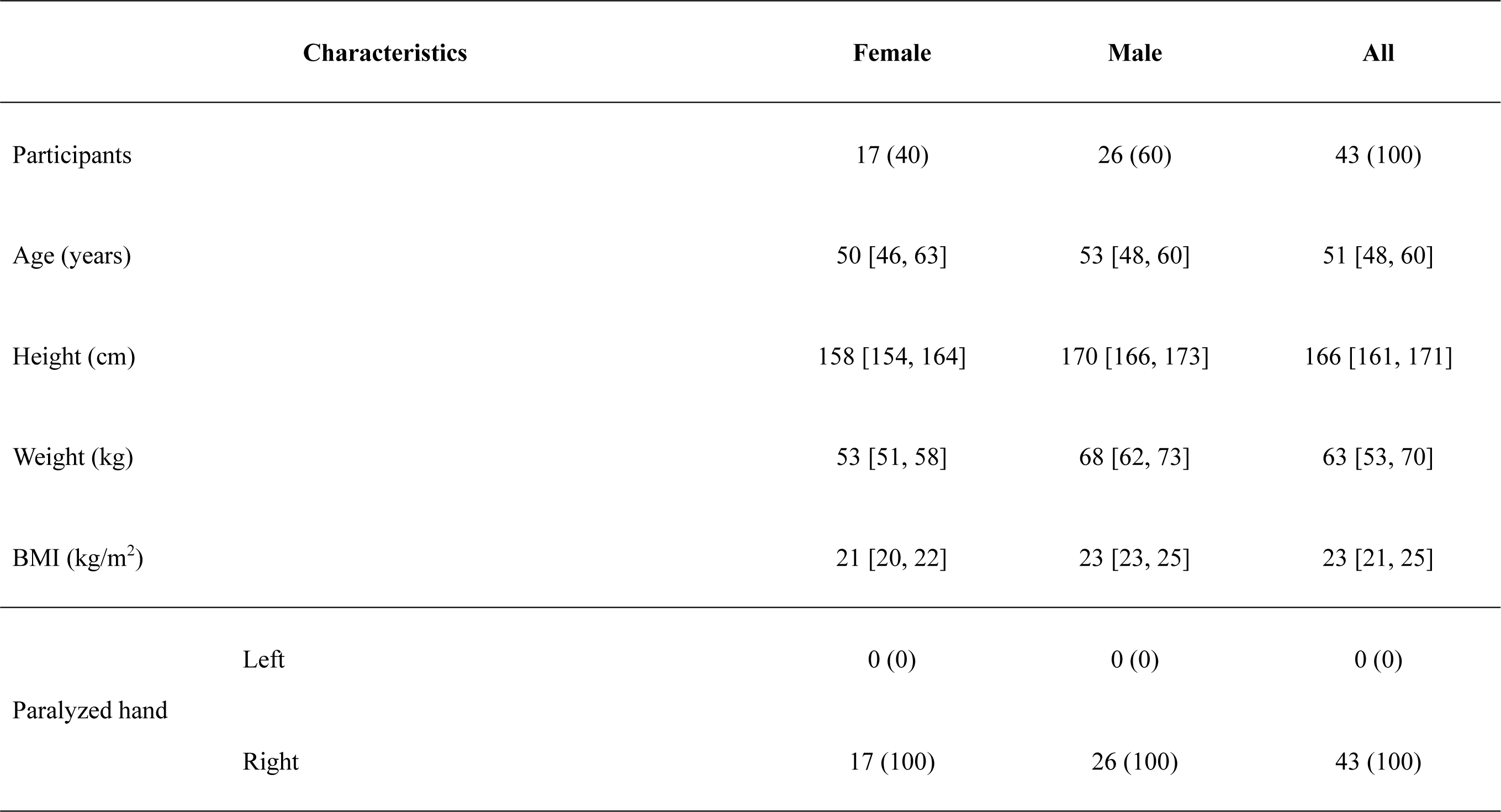
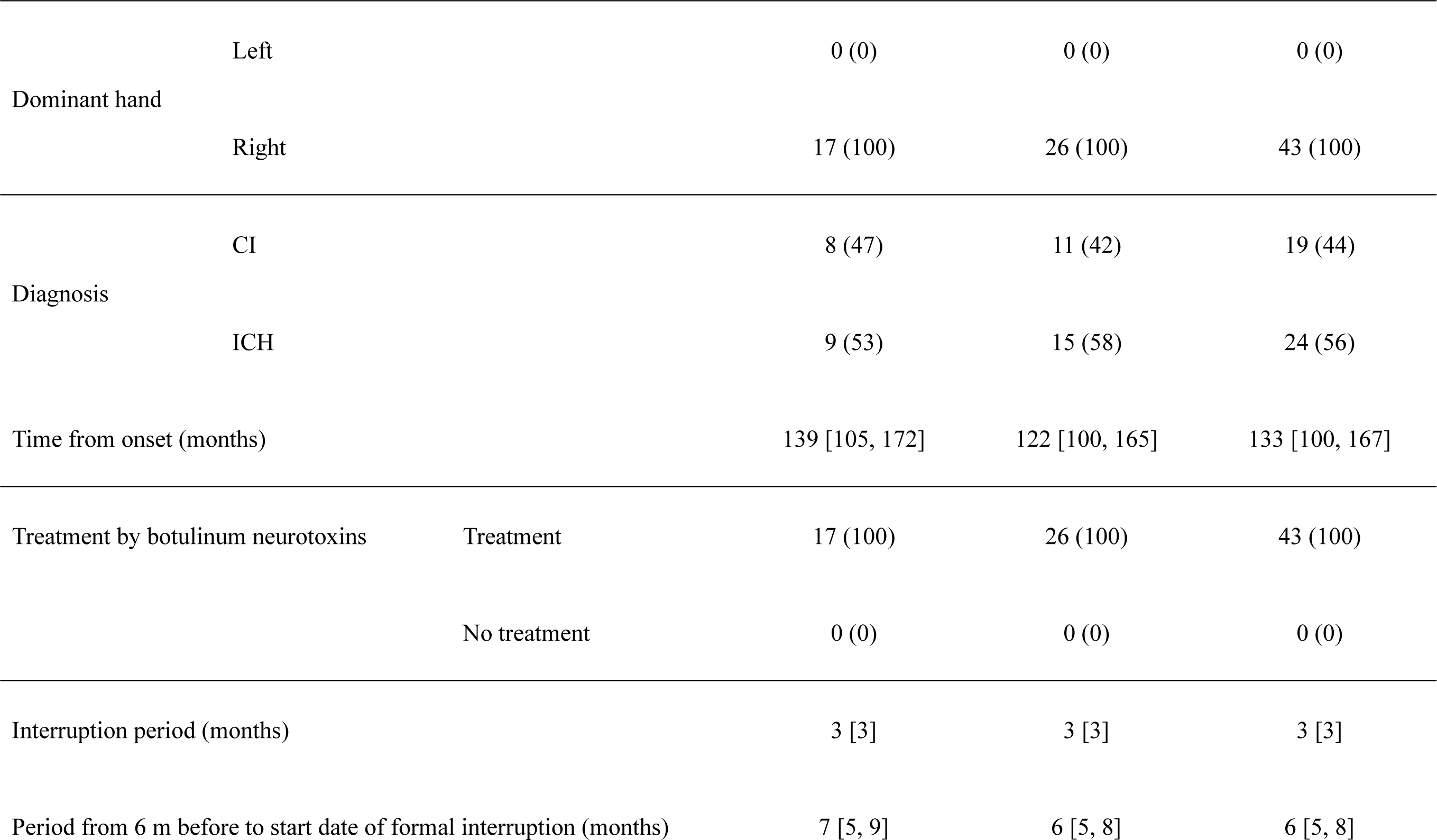
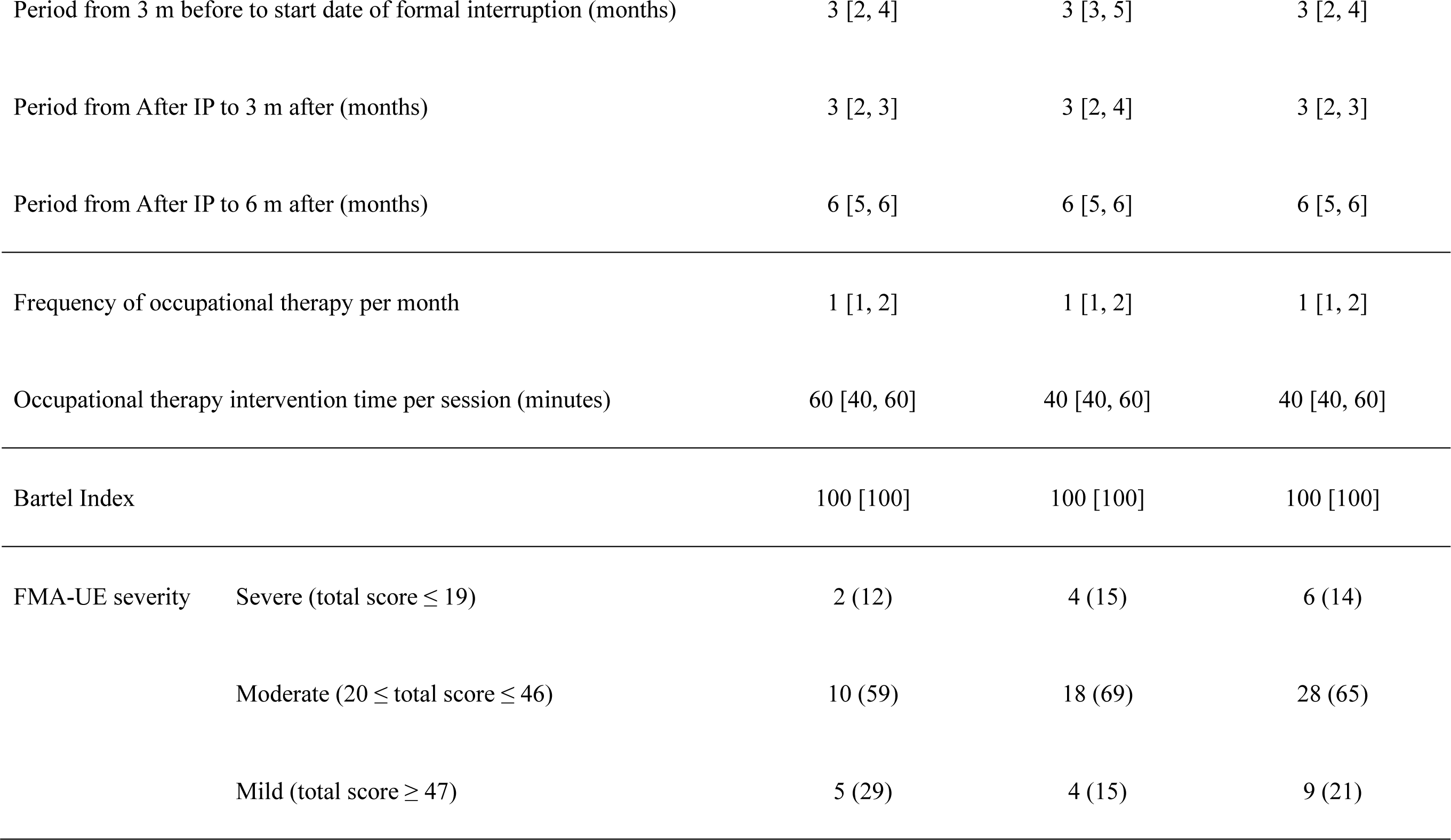
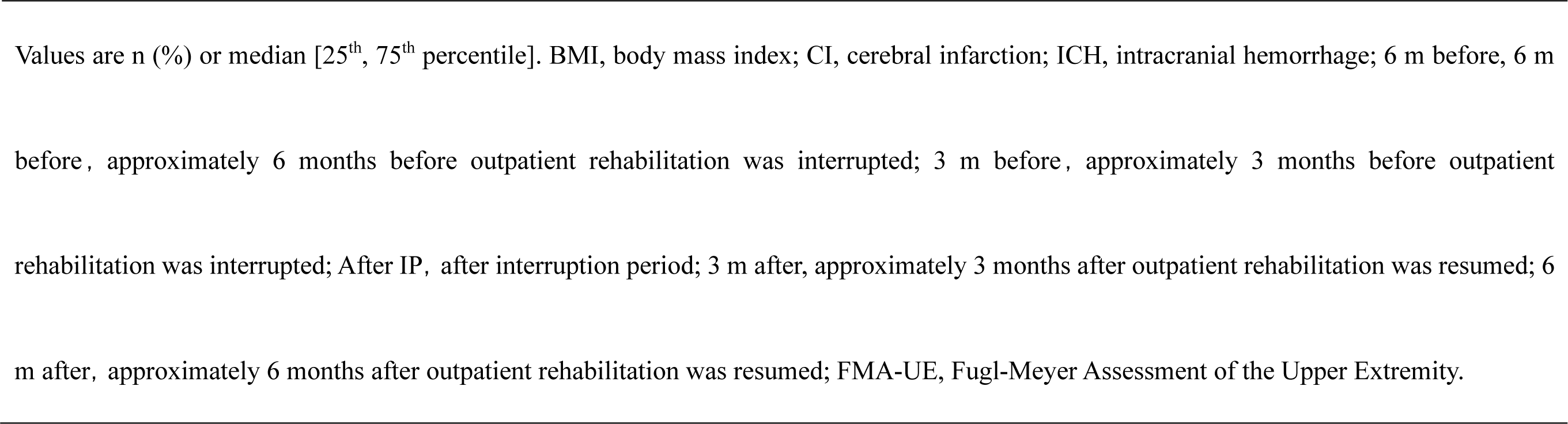
Characteristics of Analyzed Patients.

**Table 3.**
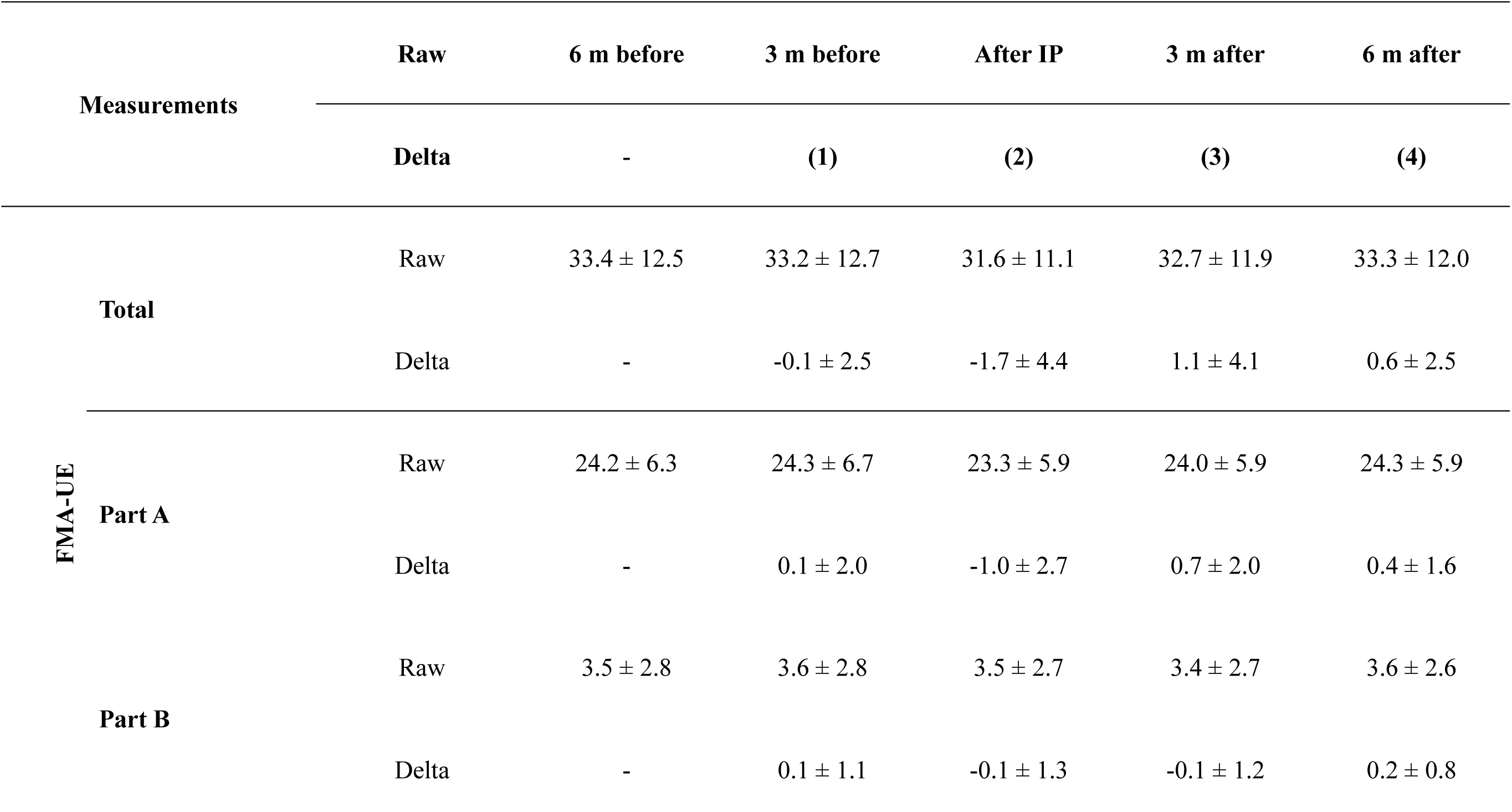
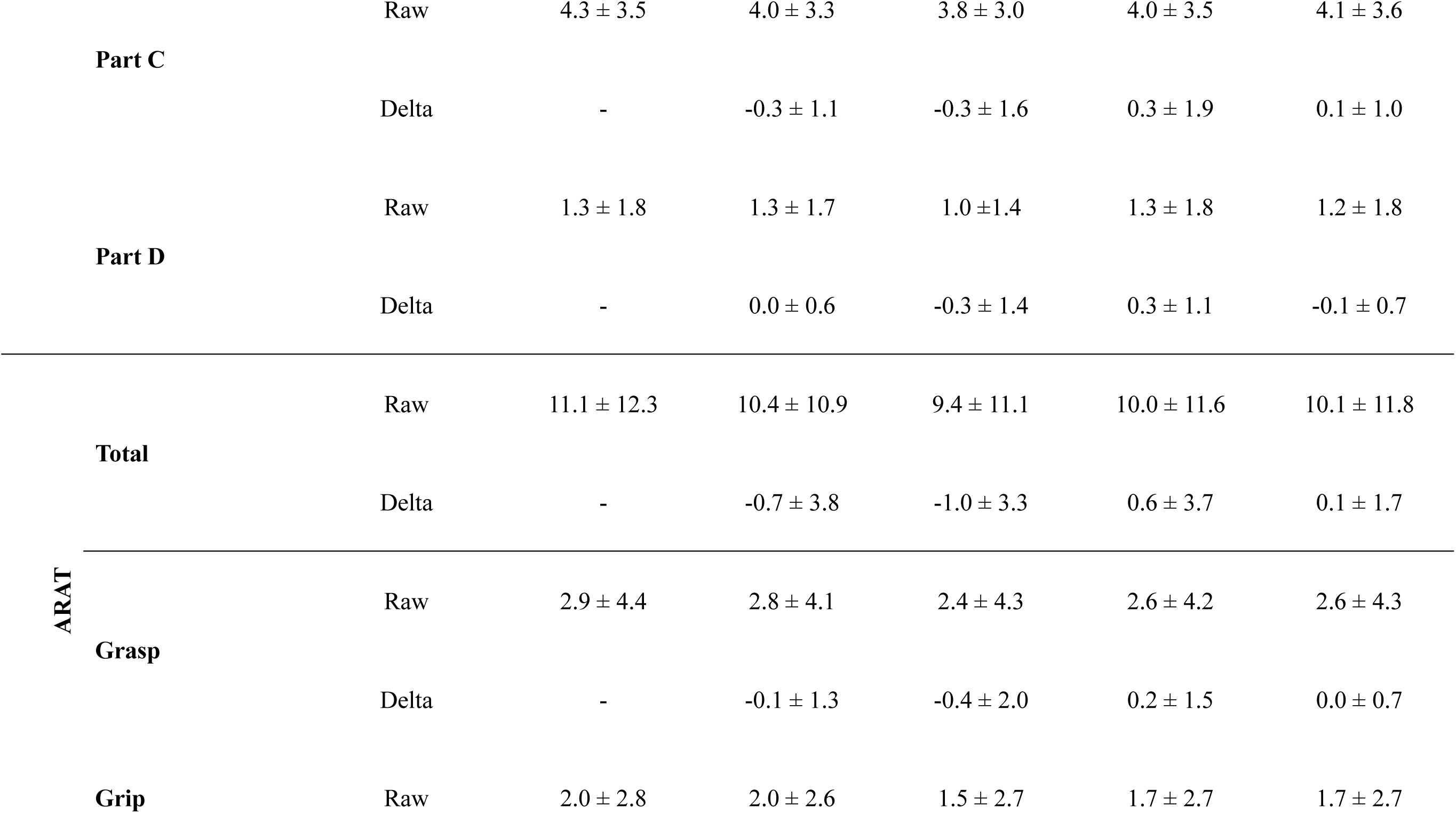
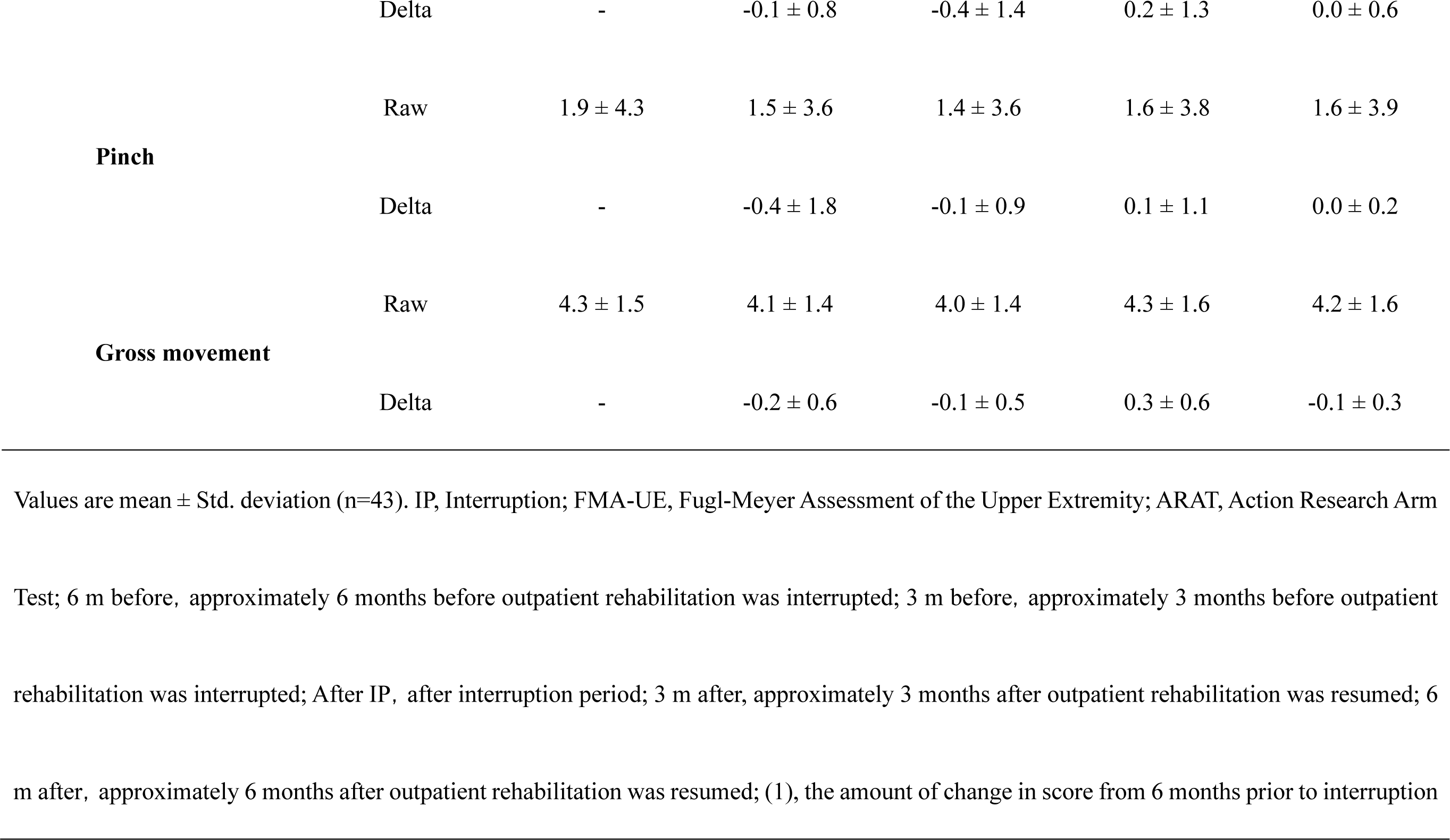
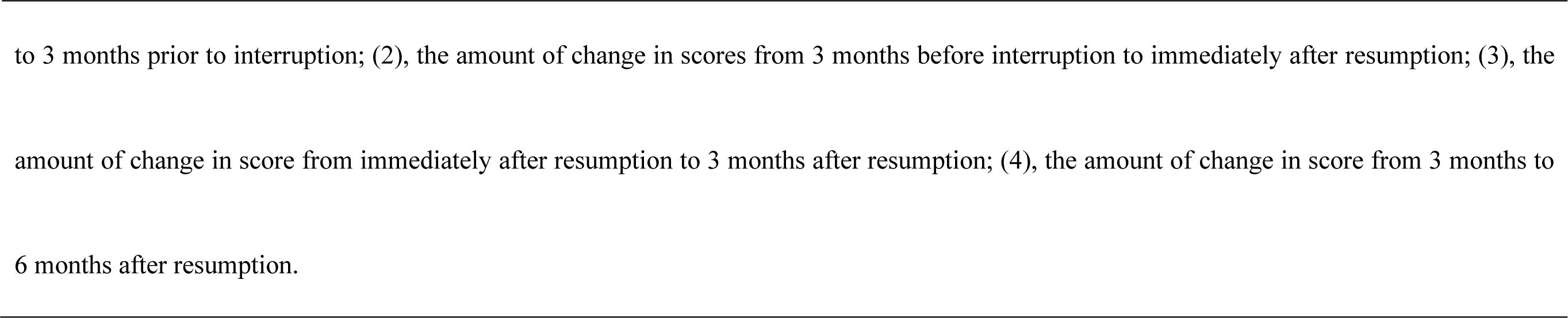
Raw and delta scores of FMA-UE and ARAT during the study period.

An analysis of covariance with the time of evaluation as a factor and the change in FMA-UE and ARAT scores as the dependent variable showed that the total (F=6.925, *p*<0.001, η^2^=0.109), part C (F=8.458, *p*<0.001, η^2^=0.131), and part D (F=11.903, *p*<0.001, η^2^ = 0.178) of FMA-UE, total (F=5.378, p=0.001, η^2^=0.116), grip (F=4.357, *p*=0.006, η^2^=0.074), pinch (F=11.685, *p*<0.001, η^2^=0.176), and gross movement (F=7.511, p<0.001, η^2^=0.116) of ARAT showed a significant main effect of time of evaluation. There was no main effect of evaluation period for part A (F=2.133, *p*=0.098, η^2^=0.037), and part B (F=1.867, *p*=0.137, η^2^=0.032) of FMA-UE, grasp (F=1.192, *p*=0.315, η^2^=0.021) of ARAT.

ANCOVA showed a significant main effect, and Tukey’s post-hoc test was performed for the items for which a significant main effect was found. In total of FMA-UE, the change(3) from immediately after resumption to 3 months after resumption was significantly higher than the change (2) from 3 months before interruption to immediately after resumption (Mean difference=2.79, 95% CI Lower=0.92, Upper =4.66, SE=0.72, t=3.88, Cohen’s d=0.66, *p*<0.001, Fig 3a). Similarly, the change (4) from 3 to 6 months after resumption was significantly higher (Mean difference=2.28, 95% CI Lower=0.41, Upper=4.15, SE=0.72, t=3.17, Cohen’s d=0.64, *p*=0.010, Fig 3a). In part D (coordination/speed) of FMA-UE, the change (3) from immediately after resumption to 3 months after resumption was significantly higher than the change (2) from 3 months before to immediately after suspension (Mean difference=0.51, 95% CI Lower=0. 01, Upper=1.01, SE=0.19, t=2.65, Cohen’s d=0.42, *p*=0.043, Fig 3b). In grip of ARAT, the change (3) from immediately after resumption to 3 months after resumption was significantly higher than the change (2) from 3 months before to immediately after suspension (Mean difference=0.61, 95% CI Lower=0.02, Upper=1.20, SE=0.23, t=2.66, Cohen’s d=0.45, *p*=0.042, Fig 3c). In gross movement of ARAT, the change (3) from immediately after resumption to 3 months after resumption was significantly higher than the change (1) from 6 months before interruption to 3 months before interruption (Mean difference=0.46, 95% CI Lower=0.18, Upper=0.75, SE=0.11, t=4.18, Cohen’s d=0.74, *p*<0.001, Fig 3d). Similarly, it was significantly higher than the change (2) from 3 months before interruption to immediately after resumption (Mean difference=0.40, 95% CI Lower=0.11, Upper=0.69, SE=0.11, t=3.55, Cohen’s d=0.67, *p*=0.003, Fig 3d). The change (4) from 3 to 6 months after re-opening was significantly lower than the change (3) from immediately after reopening to 3 months after reopening (Mean difference=-0.33, 95% CI Lower=-0.62, Upper=-0.04, SE=0.11, t=-2.92, Cohen’s d=-0.66, *p*=0.021, Fig. 3d). There were no significant differences in part C scores of FMA-UE and the total and pinch scores of ARAT by the time of evaluation.

**Fig 3.**
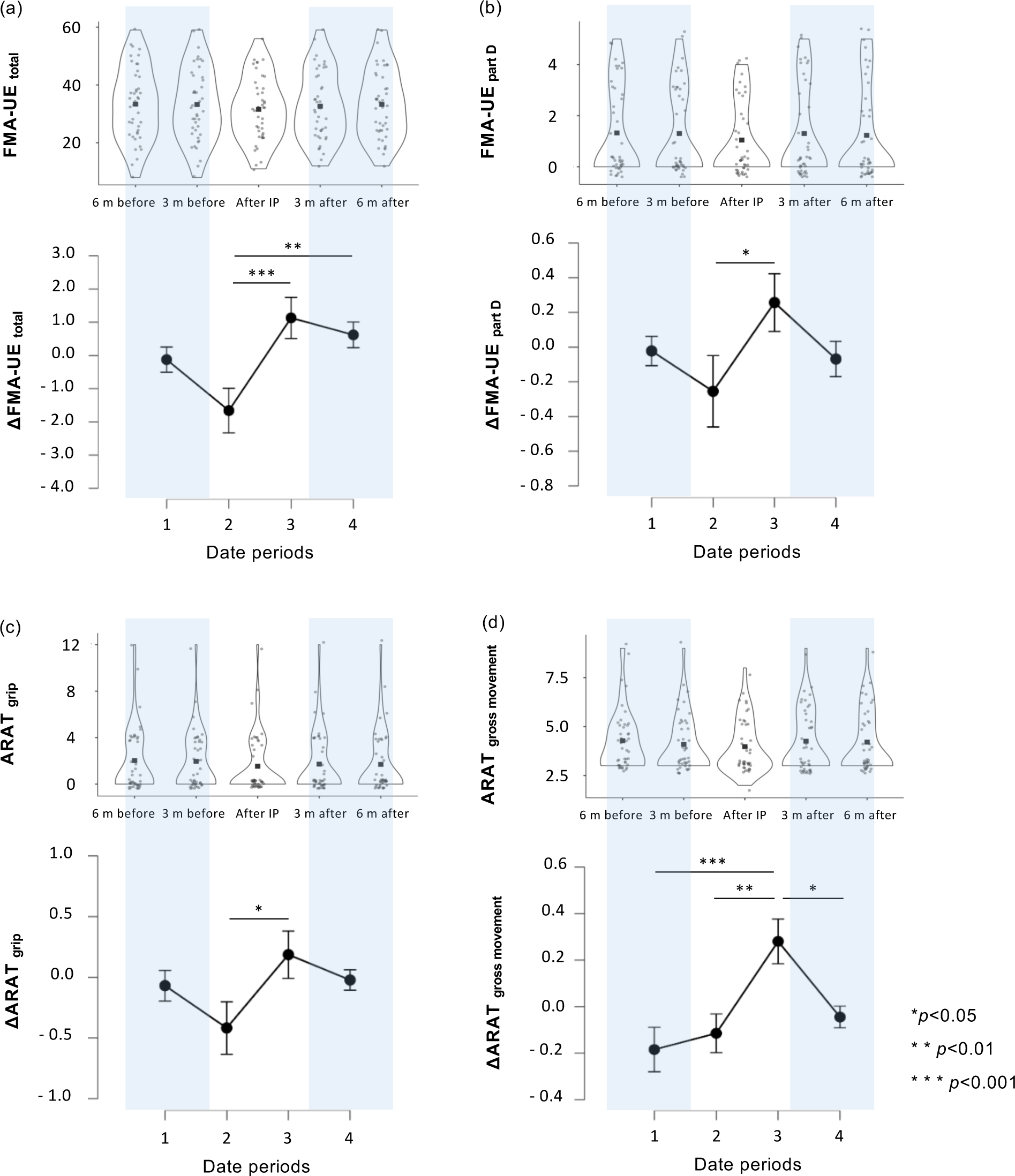
Changes in upper limb motor function over time. Scores at five time points are shown in the upper panel and changes in scores in the lower panel for (a) FMA-UE total, (b) FMA-UE part D, (c) ARAT grip, and (d) ARAT gross movement. Violin plots are displayed by a karnel density estimator, shape at each value of the x-axis represents the density of data points at that score. The black squares in the graph indicate the mean. The gray circles indicate the values for each patient. Statistical significance was set at *p*<0.05 for Tukey’s multiple comparisons (n=43). FMA-UE, Fugl-Meyer assessment of the upper extremity; ARAT, Action Research Arm Test; 6 m before, approximately 6 months before outpatient rehabilitation was interrupted; 3 m before, approximately 3 months before outpatient rehabilitation was interrupted; After IP, after interruption period; 3 m after, approximately 3 months after outpatient rehabilitation was resumed; 6 m after, approximately 6 months after outpatient rehabilitation was resumed; Date period 1, from 6 months before interruption to 3 months before interruption; Date period 2, from 3 months before interruption to immediately after resumption; Date period 3, from immediately after resumption to 3 months after resumption; Date period 4, from 3 months after resumption to 6 months after resumption.

## Discussion

This study showed that chronic stroke patients with hemiplegia where upper limb function had declined, due to the interruption of outpatient rehabilitation and refraining from going out due to the SARS-CoV-2 infection outbreak, recovered their upper limb function scores 3 months after rehabilitation was resumed. We have previously reported that upper limb function deteriorates and subjective symptoms such as joint stiffness, muscle stiffness, and muscle weakness occur in chronic stroke patients after 3 months of outpatient rehabilitation and refraining from going out [8]. Our present finding that these patients regained upper limb function after rehabilitation suggests that the effects of temporary physical inactivity can be reversed in about 3 months, if appropriate rehabilitation is resumed. Others have shown that motor function of the paralyzed upper extremity is preserved over time with sustained training [22, 23]. However, in rats with motor cortex damage, performance declines when training is interrupted, and additional training has been shown to be required to return performance to pre-interruption levels [24]. Similarly, for stroke patients, an interruption of rehabilitation generates a decline in upper limb motor function [25]. The results of the present study reaffirm the importance of continuous rehabilitation for patients and therapists.

It has been assumed that during the approximately 3 month period when outpatient rehabilitation was interrupted and patients refrained from going out, the amount of movement of the paralyzed upper limb in daily life decreased. It has been reported that the spread of novel coronavirus infection decreased the amount of physical activity in people [26, 27]. Chronic stroke patients receive outpatient rehabilitation one to four times a month. Patients go out for hospital visits and engage in occupational therapy and exercise therapy, resulting in higher physical activity. Although the amount of physical activity varies from person to person, refraining from going out to prevent SARS-CoV-2 infection is thought to have reduced the amount of activity in chronic stroke patients, causing disuse of limbs. In healthy elderly subjects, 10 days of complete rest results in a 30% decrease in muscle protein synthesis and a 16% decrease in muscle strength [28]. Since it has been reported that stroke patients have less muscle mass than healthy subjects and paralyzed side limbs of stroke patients have less muscle mass than non-paralyzed side limbs [29–31], it is inferred that motor paralysis and decreased activity have a combined impact on the limbs of the paralyzed side in stroke patients, leading to progressive disuse. Brain and nerve changes are due to disuse-dependent plasticity [32]. Disuse syndrome in stroke patients includes short-term effects on the brain and nervous system and slowly progressive effects on the musculoskeletal system over time. The patients in this study had experienced a chronic stroke, and inactivity during the period of interrupted rehabilitation was assumed to have caused a transient decline in upper limb motor function, affecting both the brain and the motor system.

The results of the present study suggest that the decline in upper limb motor function due to the interruption of outpatient rehabilitation is a reversible phenomenon. The recovery of function after 3 months of inactivity is attributed to the fact that the interview, assessment, and intervention procedures performed on the patients, within the usual scope of care, promoted use-dependent plasticity in patients. Use-dependent plasticity is a phenomenon in which repeated activation of certain neurons facilitates the same pattern of activity. Previous studies have shown that providing repetitive exercises and increasing the use of the paralyzed upper limb in daily life are intended to increase this use-dependent plasticity [32]. The amount of sufficient practice needed to restore motor paralysis is estimated to be at least 20 hours over a 2-week period of intensive practice, and at least 60 hours of practice is needed to further enhance the use of the paralyzed upper extremity [33–35]. The participants in this study received approximately three sessions of outpatient rehabilitation during the first 3 months after resumption of medical care, with each session lasting approximately 40 minutes. The amount of time the patients spent practicing independently increased under the guidance of the therapists, and the amount of time spent practicing reached the amount of practice required to recover from motor paralysis, suggesting that the upper limb motor functions had been restored. This result confirms that if therapists can increase the amount of physical exercise for patients, they can prevent functional decline and promote functional recovery, even with limited intervention opportunities. Recovery of motor paralysis is facilitated when patients are aware of their own goals and carry out physical exercises independently, and when they are given feedback by the therapist [36, 37]. In the course of a typical rehabilitation management of a patient, the resolution of issues that arise in the patient are shared with the patient and the therapist, and the treatment goals and practice plan are agreed upon and provided to the patient. During the current study period, the therapists provided feedback to the patients in the form of brief, easy to remember verbal feedback such as “work together to achieve the goal,” “a sufficient amount of practice will promote recovery,” “better use of the disabled hand will make ADL easier.” These procedures, which were implemented after the patient resumed outpatient care, motivated the patient to engage in functional practice and to use the paralyzed upper extremity in ADL, and this increased the amount of movement of the paralyzed upper extremity, and motor function recovered.

Treatment goal setting, amount of practice, and patient motivation are important for recovery of motor paralysis after stroke [38–40]. Intervention outcomes improve when patients are allowed to monitor their own condition and are motivated to practice [41]. The patient upper extremity function, amount of use of the paralyzed upper extremity, and self-training were assessed at the time of resumption of outpatient rehabilitation within the usual scope of practice. The results of these assessments were compared with those obtained by the occupational therapists in their previous practice to clarify the changes that had occurred in the patient and to establish treatment goals and exercises. The assessments made after the resumption of outpatient rehabilitation were shared individually with the therapists and patients, and the patients were made aware of the changes that had occurred in their physical function and their future treatment goals. This was assumed to motivate the patients to engage in the recommended physical exercises.

When the therapist encourages the patient to exercise, information about the loss from not exercising and the gain from carrying it out impacts the patient according to the personality, psychological state, and social circumstances of the patient [42, 43]. During intervention procedures for resuming activities in stroke patients who are inactive or were temporarily inactive, therapists can use our data for patients as follows. If the patient does not practice recommended activities voluntarily or does not increase the use of his/her hands in daily life, it can be used as information about the loss of motor function and the risk of further deterioration of ADLs. If the patient hesitates in carrying out physical activities when rehabilitation can be resumed, the same period of inactivity can be presented as information about gains that can facilitate recovery by intensifying practice aimed at improving decreased function and movement and increasing the use of the paralyzed upper extremity in ADL. For patients with family members or caregivers, this can be used as the basis for suggestions such as, “by using your hands better, you can reduce the burden on your family and help caregivers back to a normal routine.” In this study, recovery of upper limb function was suggested in part D scores of the FMA-UE, and grip and gross movement scores of the ARAT. These subtests assess smoothness and speed of movement in part D of the FMA-UE, manipulation of objects including forearm rotation in the ARAT grip, and reaching movement in changing, dressing, and bathing activities in the gross movement [14, 17]. During outpatient rehabilitation, occupational therapists instructed the patients in ameliorating decreased upper limb function and encouraged the use of the paralyzed upper limb in a sitting position and in ADL at home. The occupational therapists considered the severity of motor paralysis in patients. The motor components that improve in patients with chronic stroke differ according to the severity of motor paralysis, even when assessed during a 2-week treatment combining transcranial magnetic stimulation and occupational therapy [44]. Patients with more severe motor paralysis showed greater recovery in ARAT grip and gross movement scores, while those with less severe motor paralysis were more likely to improve the quality of life activities using the paralyzed upper extremity. Reports examining the degree of difficulty in the sub-items of the FMA-UE have shown that the items in part D are more difficult [45, 46]. In patients with mild disease, occupational therapists facilitated the use of the paralyzed upper limb for ADL, and as a result, the smoothness and speed of movement in part D were restored. On the other hand, in patients with severe disease, the range of reach of the paralyzed upper limb was increased by exercises to improve the joint range of motion and muscle flexibility of the paralyzed upper limb, by exercises focused on ADL. As the opportunity to use the paralyzed upper limb as an aid in sitting movements and to manipulate objects on a desk increased, so did ARAT scores for grasp and gross movement improve.

This study documents the decline in upper limb function and the process of functional recovery in patients with chronic stroke after 3 months of interrupted outpatient rehabilitation. The results of this study can be used in the future to help prevent functional decline in patients who are restricted from leaving their homes due to the spread of various infectious diseases, and to devise ways to help patients recover after limb motor function decline due to interrupted rehabilitation. In this study, occupational therapists interviewed patients about their subjective symptoms and ADL that occurred during the 3 month interruption period, conducted an objective functional assessment, and identified problems occurring in the patients. They provided treatment and guidance to the patients and promoted motivation. The amount of physical activity and use of the paralyzed upper extremity can be evaluated by using pedometers, activity meters, or by measuring the amount of movement with a device attached to the paralyzed upper extremity [47, 48]. These methods may be effective tools for monitoring and providing feedback when assisting patients who have opted to refrain from leaving their homes due to telerehabilitation. In order to maintain and improve patient motivation for rehabilitation, it is important for therapists to set clear goals, such as using the Canadian Occupational Performance Measure [49] or the Aid for Decision-making in Occupation Choice [50]. For patients who require Botulinum toxin treatment for spasticity, such as the participants in this study, the introduction of low-frequency therapy devices or vibration stimulation devices may also be considered as devices to assist independent practice. It has been reported that these therapeutic devices can enhance the therapeutic effect when used in conjunction with regular rehabilitation [51, 52]. The therapist is required to evaluate these therapeutic devices and the subjective physical symptoms of the patient, prescribe appropriate physical activity practices, and suggest the introduction of appropriate devices for the patient as an adjunctive means of treatment.

This study has several limitations. First, the amount of physical activity of patients during the period when rehabilitation was interrupted and the amount and duration of self-training after rehabilitation resumed were not investigated. The amount of activity is a factor that influences the maintenance of physical function, and we hypothesized that refraining from going out to prevent SARS-CoV-2 infection was a behavior that decreased the amount of activity in patients with chronic stroke. The extent to which the activity level of chronic stroke patients changed during and after the interruption of outpatient rehabilitation should be investigated when designing new studies in the future. Second, this study did not examine the nutritional status and sleep duration of the patients. These are factors that affect the neuroplasticity of the brain, and this provides the neural basis of use-dependent plasticity [53]. Third, because the participants in this study were generally independent in their daily lives, it is not possible to use the results of this study to estimate the amount of recovery of upper limb function in the scenario of severely ill patients who require assistance in daily activities. Finally, because this study was conducted at a university hospital in Tokyo, Japan, there are limitations to the generalizability of the study in estimating the amount of recovery of upper limb function for patients residing in other geographical regions where access to public transportation and the extent of hospital facilities may be very different from this patient cohort.

## Conclusion

The results of this study suggest that the loss of upper limb function in chronic stroke patients caused by the restriction of outings and interruption of outpatient care due to the spread of SARS-CoV-2 infection was reversible, and that upper limb function was restored 3 months after the resumption of rehabilitation. The temporary functional losses incurred by the patient and the subsequent recovery values can serve as reference standards for the rehabilitation of a patient who has had a period of inactivity of about 3 months.

## Data Availability

Data Access / Ethics Committee (contact via) for researchers who meet the criteria for access to confidential data.

## Acknowledgments

We would like to thank all occupational therapists of the Department of Rehabilitation, Jikei University School of Medicine, for their cooperation in obtaining operational approval and data for conducting this study.

## Notes

### Competing Interest Statement

The authors have declared no competing interest.

### Funding Statement

The funders had no role in study design, data collection and analysis, decision to publish, or preparation of the manuscript.

### Author Declarations

The study was approved by the Ethics Committee of the Jikei University School of Medicine (Approval Number 24-295-7061).

